# Patient-Reported Food Noise, Cravings and Appetite in Adults with Obesity Treated with Weekly Injectable GLP-1 Based Therapies: A Prospective Real-World Study

**DOI:** 10.64898/2026.11.13.26367932

**Authors:** Carel Le Roux, Bethany Kalich, Chalak Muhammad, Deborah Horn, Kate Orroth, Leandro Boer, Min-Jung Wang, Abd A Tahrani, Suna Avcil, Lam Nguyen, Zheng-Rong Li, Stephen Fabiano, C Michael Gibson, Yu Mi Kang

## Abstract

**Aims:** Although pivotal trials have established efficacy in weight reduction during the use of GLP-1 based therapies, daily fluctuations in food-related experiences between scheduled assessments remain to be better characterized. We therefore aimed to assess day-to-day patterns in food noise, cravings, hunger, satiety, and fullness among adults receiving weekly semaglutide or tirzepatide for weight management.

**Materials and Methods:** This prospective, direct-to-participant, US observational study enrolled adults receiving weekly subcutaneous semaglutide or tirzepatide for weight management with self-reported BMI ≥30 kg/m² before treatment initiation. Participants completed daily 24-hour-recall electronic diaries assessing food-related experiences for 6 weeks. Each injection defined Day 1 of a new dosing cycle.

**Results:** Of 275 enrolled participants, 237 (86%) were included in the analysis. Daily food noise prevalence ranged from 4.3% to 23.8% over 6 weeks, generally highest on dosing day, declining thereafter, and returning toward dosing-day levels by Day 7. Overall, 182 participants (76.8%) reported food cravings at least once over 6 weeks, with moderate craving intensity as most frequently reported (98/237 [41.4%]). Across the study period, mean visual analog scale (0-100) on cycle Days 1, 2, and 7 were 39.9, 31.1, and 41.6 for hunger; 66.3, 69.8, and 64.2 for satiety; and 62.0, 65.1, and 60.6 for fullness.

**Conclusions:** Food noise, cravings, and hunger declined soon after GLP-1 based therapy administration and increased towards the end of the weekly interval, whereas fullness and satiety fluctuated less. Daily assessments may capture experiences missed by conventional trial visit schedules and may be particularly relevant as therapies with longer half-lives become available.

## Introduction

Glucagon-like peptide-1 (GLP-1)-based therapies have emerged as effective treatments for excess body weight^1, 2^ and associated cardiometabolic risk.^3^ Beyond their effects on weight and weight-related outcomes, there is growing interest in how these therapies may influence food-related experiences.

Emerging evidence suggests that GLP-1-based therapies may improve subjective food-related experiences, including food noise, cravings, hunger, satiety, and fullness, associated with GLP-1-based therapies.^4, 5^ In a 6-week study including adults without diabetes and with a BMI of 27–50 kg/m², tirzepatide (5 mg weekly for 3 weeks, followed by 10 mg weekly for 3 weeks) reduced food cravings, perceived hunger, and other measures of food behavior, compared with placebo.^4^ In the STEP 5 trial, exploratory assessments using the Control of Eating Ǫuestionnaire (CoEǪ) demonstrated improvements in several craving-related outcomes with semaglutide (2.4 mg) at week 20, although these measures were no longer different from baseline during the weight-stabilization phase at week 52, and 104.^5^ Collectively, these findings suggest favorable effects of GLP-1-based therapies on food-related experiences, particularly during the active weight reduction. However, the earliest follow-up assessments in these studies occurred 5 to 20 weeks after baseline, potentially overlooking day-to-day fluctuations in food-related experiences, particularly during treatment initiation and across weekly dosing cycles. Characterizing these daily experiences may provide a more granular understanding of patient-reported treatment effects and inform future investigations into their potential relationships with treatment satisfaction, adherence, and clinical outcomes.

An additional challenge is the lack of standardized, well-validated instruments that consistently distinguish among different domains of food-related experiences. For example, food noise and food cravings are sometimes used interchangeably in surveys, although food noise generally refers to persistent or intrusive food-related thoughts, whereas cravings involve urges, often difficult to resist, to consume specific foods.^6, 7^ Similarly, satiety (lower appetite after eating, which sets the interval before the next meal) and fullness (the physical sensation of gastric filling)^8^ are not always clearly differentiated in assessment instruments.^9^ These distinctions are clinically relevant, as individuals with obesity may experience different predominant patterns of food-related behavior, potentially contributing to heterogeneity in their treatment experiences. More precise characterization of these domains may ultimately help identify patient-specific treatment effects and inform individualized therapeutic approaches.

Therefore, in this prospective, US-based observational study, daily electronic diary (eDiary) assessments were used to characterize food noise, food cravings and craving intensity, hunger, satiety, and fullness over 6 weeks among adults with obesity receiving weekly injectable GLP-1 based therapy.

## Materials and Methods

### Study design and participants

This was a prospective, US-based observational real-world study of adults with obesity receiving weekly injectable GLP-1 based therapy marketed at the time of the study for the purpose of weight management (i.e., semaglutide or tirzepatide). Participants were followed remotely for 6 weeks and completed daily electronic patient-reported outcome (ePRO) assessments using a 24-hour recall period.

At screening, participants completed a questionnaire collecting demographic, socioeconomic and clinical information, including age, sex, race and ethnicity, education, employment, household income, insurance status, medical history, obesity-treatment history, current GLP-1 based therapy, dose, categorical duration of use, and intention to continue therapy during the study. This requirement enabled assessment of food-related experiences during ongoing weekly therapy.

Participants reported their treatment status at screening. After consent and identity verification, medical records were reviewed for GLP-1 product, dose, and treatment start date to confirm eligibility and cohort assignment. When self-reported information conflicted with the medical record, the record was used when sufficient documentation was available. Participants were then classified into two cohorts based on their self-reported treatment status and the medical record findings at screening. Cohort A included participants who had received therapy for 1 to <12 weeks (early weight reduction phase) and thus, were in dose titration/escalation (tirzepatide [2.5, 5.0, 7.5, or 10 mg] or semaglutide [0.25, 0.5, or 1.0 mg]). Cohort B included participants who had received a stable dose for ≥12 weeks (late weight reduction phase), with no planned change in dose, dosing frequency, or product during the study as reported at screening (tirzepatide [5.0, 7.5, 10, 12.5, or 15 mg] or semaglutide [1.7 or 2.4 mg]). For each weekly dosing cycle, Day 1 was defined as the participant-reported injection day regardless of the day of the week and Days 2–7 as the subsequent six calendar days.

### Eligibility criteria

Adults aged ≥18 years with self-reported BMI ≥30 kg/m² before GLP-1–based therapy were eligible if they planned to continue treatment and complete daily eDiaries. Key exclusion criteria were diabetes, use of unapproved (including compounded) GLP-1 products, recent or planned use of other weight-management treatments, pregnancy or breastfeeding, and conditions or procedures that could interfere with participation; bariatric surgery within the previous year; or plans for bariatric surgery within the next 3 months. A complete list of inclusion and exclusion criteria is provided in **Supplementary Table S1.**

### Study procedures and data collection

Participants provided an Institutional Review Board (IRB)-approved electronic informed consent including authorization under the Health Insurance Portability and Accountability Act (HIPAA) to use their medical records for research purposes.

Participants were recruited remotely through Bridge Access Solutions using established recruitment channels including clinics, telehealth practices, social media, and partner gyms. Clinic and telehealth channels used chart prescreening to identify potential invitees; social media and gym channels used IRB-approved advertisements, posts, flyers, or ǪR codes.

Following informed consent and HIPAA authorization, eligibility was assessed using participant-reported information and, where available, verified against electronic medical record (EMR) data through Health Information Exchange (HIE) linkage. Before study implementation, participant-facing materials and eDiary items were assessed through cognitive debriefing with a planned sample of 7–10 adults with obesity receiving GLP-1 based therapy. This qualitative assessment evaluated the relevance and comprehensiveness of the eDiary content, and the understandability and usability of its instructions, items, and response options. Feedback and documented recommendations are summarized in **Supplementary Table S2**.

Subsequently, participants completed daily ePRO assessments remotely for 6 weeks. Each assessment used a 24-hour recall period, and participants were instructed to complete the eDiary once daily, preferably at the same time each day. Participants who missed entries received scripted telephone outreach. Reminder emails were sent after 3 consecutive missed diaries or when completion fell below 80%. The study team reviewed low-completion cases before additional outreach.

### Participant-reported outcomes and endpoints

Among the participant-reported 6-week follow-up assessments were food-related behaviors and gastrointestinal adverse effects. This analysis focused on food-related experiences, including food noise, cravings, hunger, satiety, and fullness, assessed daily over the 6-week follow-up and summarized by day of the weekly dosing cycle. Food noise and food craving presence were assessed with “yes/no” items. The eDiary also included a “yes/no” item on whether thoughts about food had negative effects on the participant or their life. When food cravings were reported, participants rated craving intensity using five response categories: very slight/mild urge, mild, moderate, severe, or very severe urge. Hunger, satiety (satisfaction), and fullness were each rated using a 0– 100 visual analog scale (VAS), with higher scores indicating greater hunger, satiety, or fullness, respectively. The eDiary items and response options are provided in **Supplementary Table S3.**

All items referred to the preceding 24 hours. Each participant-reported dose initiated a dosing cycle where Day 1 was the reported dose day, followed by Days 2–7. A cycle ended after 7 days or at the next reported dose, whichever occurred first.

### Statistical analysis

The analysis population comprised eligible participants with at least one available post-enrollment ePRO assessment. Participants who reported use of a GLP-1 based therapy in the weekly dosing cycle survey that was inconsistent with the GLP-1 based therapy reported at screening were excluded from the analysis population. For participants without linked medical records, the screening-reported therapy was used for comparison. No missing data were imputed. Outcomes were summarized descriptively for the overall analysis population using available observations. Categorical outcomes are presented as frequencies and percentages; continuous outcomes as the number of observations, mean (standard deviation [SD]), median (Ǫ1, Ǫ3), and interquartile range (IǪR; Ǫ3 minus Ǫ1), as applicable. Denominators varied by outcome and time point according to data availability. Comparisons across dosing-cycle days are descriptive.

For dosing-cycle analyses, participants reporting dose administration on consecutive dose days were excluded; other reported intervals shorter or longer than 7 days were retained according to the reported injection dates. When a reported interval could reflect an erroneous eDiary entry, participants were contacted by phone or email during a 10-day clarification period. Dosing variability was defined as at least one confirmed interval between reported doses that was not 7 days.

## Results

Of 2311 individuals who initiated the screening process, 1662 did not complete screening or did not meet eligibility criteria, and the remaining 649 participants proceeded to EMR verification. A total of 275 participants who passed EMR verification were enrolled. Mean (SD) age was 49.3 (11.7) years; 192 (69.8%) participants identified as female, and 207 (75.3%) identified as White or Caucasian. Tirzepatide use was reported by 230 participants (83.6%) and semaglutide by 45 (16.4%). No product-specific quota or restriction was applied, and all recruitment channels used the same eligibility criteria and had access to users of either product. While all the participants had BMI ≥30 kg/m^2^ before initiating GLP-1 based therapy, mean (SD) BMI at enrollment was 32.2 (7.5) kg/m²; 121 participants (44.0%) had an enrollment BMI <30 kg/m² (**Table 1**). A total of 242 participants completed at least one post-enrollment eDiary assessment. Of these, 237 were included in the analysis population; five were excluded because the GLP-1 based therapy reported during the study was inconsistent with screening and/or available medical-record data (**Figure 1**). A total of 155 participants (65.4% of the 237 participants in the analysis population) provided an eDiary entry at the start of Week 6. The number of available entries declined over the study period. A total of 40 participants provided an entry on Week 6 Day 7 (17.5% of the 237 participants in the analysis population).

**Table 1:** Baseline demographic and treatment characteristics of enrolled participants.

| **Characteristic** | **Overall population (N=275)** |
|---|---|
| **Mean (SD) age, years** | 49.3 (11.7) |
| **Sex, n (%)** |  |
| Female | 192 (69.8) |
| Male | 82 (29.8) |
| Prefer not to answer | 1 (0.4) |
| **Race and ethnicity, n (%)** |  |
| White or Caucasian | 207 (75.3) |
| Black or African American (non-Hispanic) | 21 (7.6) |
| Latino or Hispanic | 11 (4.0) |
| Asian or Asian American | 6 (2.2) |
| Other, multiple races, prefer not to answer, or missing | 30 (10.9) |
| **BMI, kg/m<sup>2</sup>** |  |
| Mean (SD) | 32.2 (7.5) |
| Median (Q1 and Q3) | 31.2 (26.6-36.3) |
| <30 kg/m<sup>2</sup>, n (%) | 121 (44.0) |
| 30 to <35 kg/m<sup>2</sup>, n (%) | 72 (26.2) |
| 35 to <40 kg/m<sup>2</sup>, n (%) | 42 (15.3) |
| ≥40 kg/m<sup>2</sup>, n (%) | 40 (14.5) |
| **Cohort at enrollment, n (%)** |  |
| Cohort A: 1 to <12 weeks of therapy | 137 (49.8) |
| Cohort B: ≥12 weeks of therapy | 138 (50.2) |
| **Weekly GLP-1 based therapy, n (%)** |  |
| Tirzepatide | 230 (83.6) |
| Semaglutide | 45 (16.4) |
| **Selected self-reported medical conditions, n (%)** |  |
| Hypertension | 106 (38.5) |
| Hypercholesterolemia | 89 (32.4) |
| Obstructive sleep apnea | 86 (31.3) |

Values are n (%) unless otherwise indicated. BMI was calculated using self-reported height and weight at enrollment. BMI before GLP-1 based therapy initiation was self-reported and used to assess eligibility; BMI at enrollment was not required to be ≥30 kg/m<sup>2</sup>. Medical conditions were self-reported; participants could report more than one condition.
BMI, body mass index; GLP-1, glucagon-like peptide-1; N, total number of participants; n, number of participants with the specified characteristic; SD, standard deviation.

**Figure 1:**
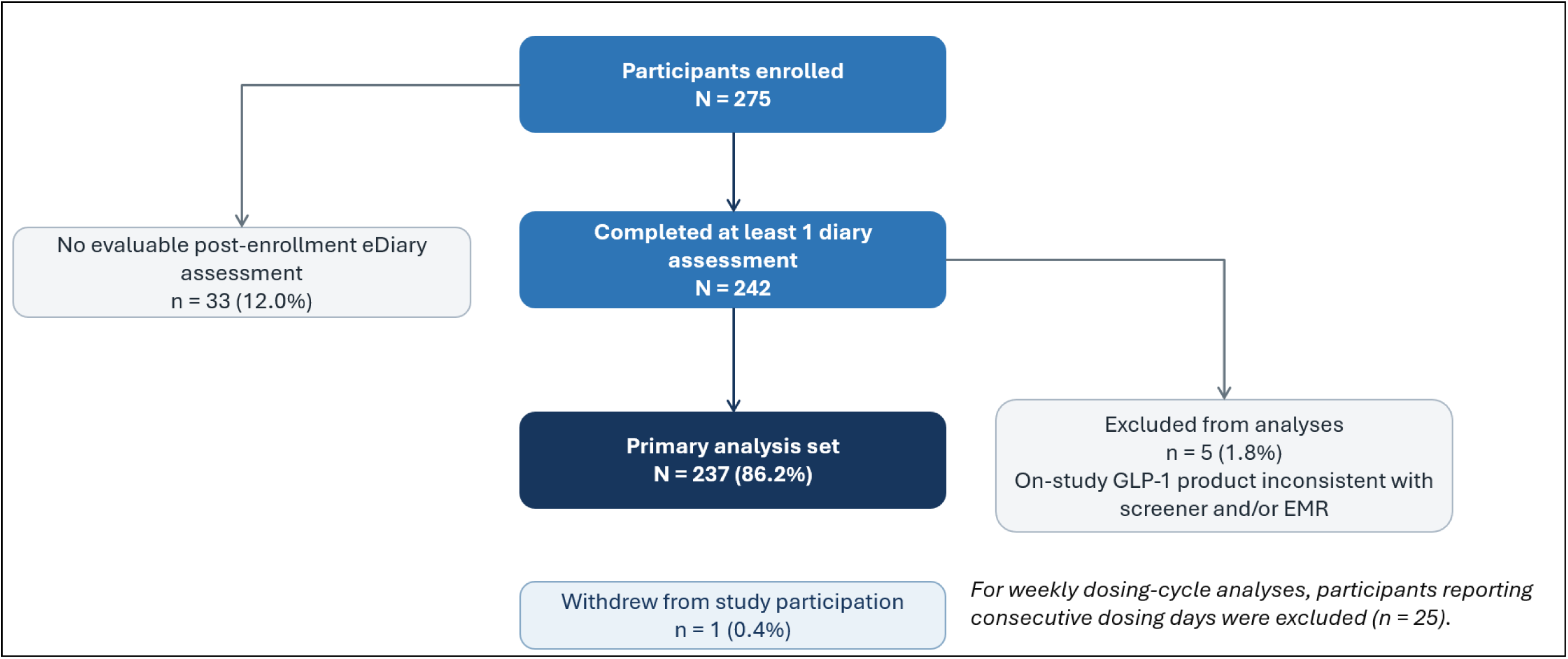
Participant enrollment, eDiary completion, and analysis populations. eDiary, electronic diary; EMR, electronic medical record; GLP-1, glucagon-like peptide-1; N, total number of participants; n, number of participants.

### Food noise prevalence and impact on participants

Within each of the six weekly dosing cycles, observed daily food noise prevalence was highest on the dosing day, decreased in the days following injection, and generally increased again toward Day 7 (**Figure 2; Supplementary Figure S1**). In Week 1, food noise prevalence was 23.8% on Day 1 (51/214), 10.1% on Day 2 (21/208), and 21.5% on

**Figure 2.**
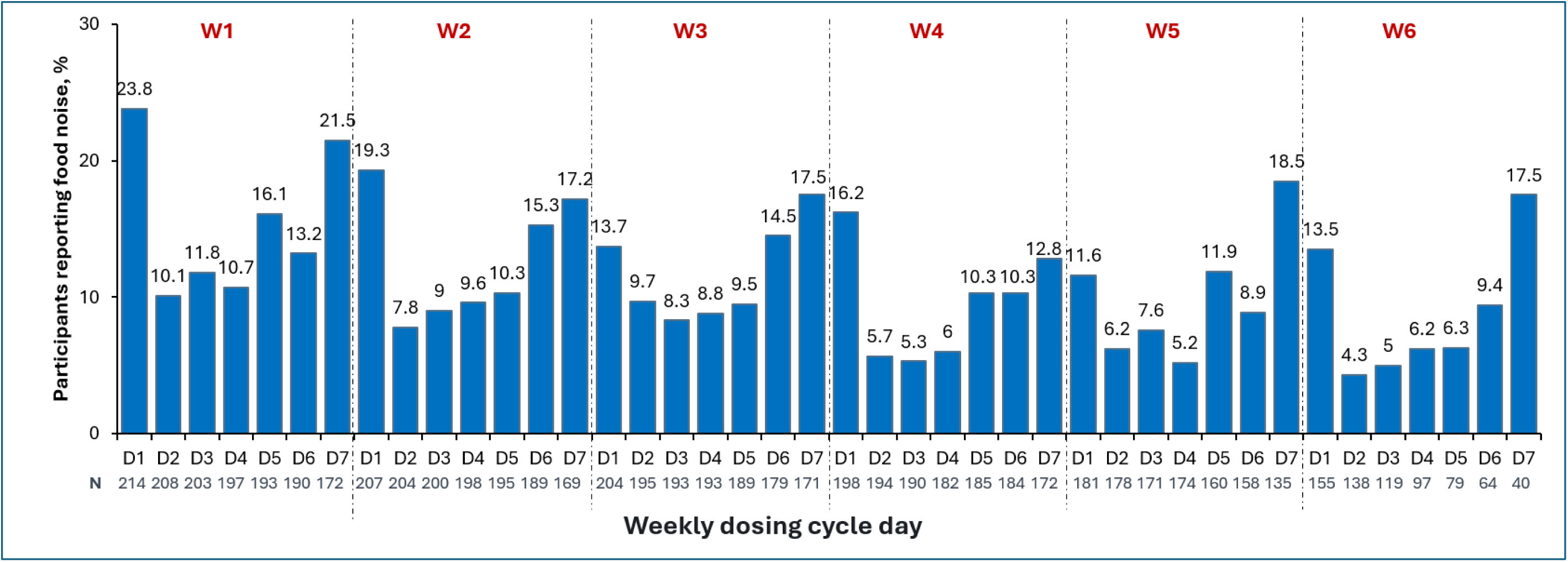
Daily food noise reports by day of the weekly dosing cycle (Primary analysis set) Values are percentages of participants with evaluable food noise data for the specified dosing-cycle day. N indicates the number of participants with evaluable data for that day. Food noise was defined for participants as unwanted, distracting, and hard-to-stop thoughts about food. Each dosing cycle began on a participant-reported dose date and included up to the next 7 days or the next reported dose, whichever occurred first. Participants reporting consecutive dosing days were excluded from cycle-aligned analyses. D, day; W, week.

Day 7 (41/190). In Week 6, prevalence was 13.5% on Day 1 (21/155), 4.3% on Day 2 (6/138), and 17.5% on Day 7 (7/40). Fewer participants had evaluable food noise data in Week 6 than in Week 1. Over the 6-week observation period, food noise was reported on a mean (SD) of 5.4 (7.7) days (**Table 2**). Negative impact of food-related thoughts on participant or life was reported by 85 of 237 participants (35.9%) at least once during the 6-week period **(Supplementary Table S4)**.

**Table 2:**
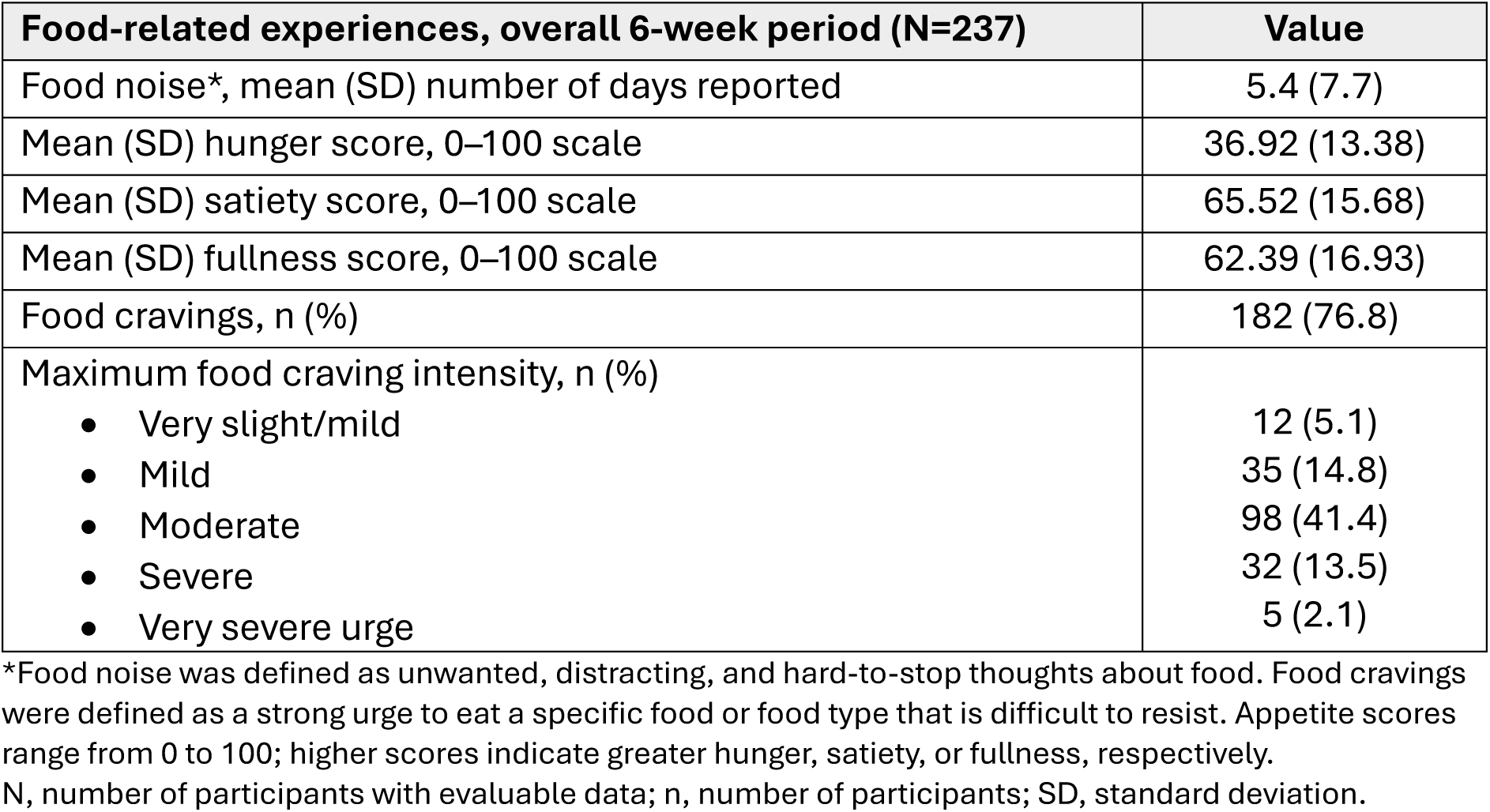
Participant-reported food-related experiences during the 6-week observation period.

| **Food-related experiences, overall 6-week period (N=237)** | **Value** |
|---|---|
| Food noise*, mean (SD) number of days reported | 5.4 (7.7) |
| Mean (SD) hunger score, 0–100 scale | 36.92 (13.38) |
| Mean (SD) satiety score, 0–100 scale | 65.52 (15.68) |
| Mean (SD) fullness score, 0–100 scale | 62.39 (16.93) |
| Food cravings, n (%) | 182 (76.8) |
| Maximum food craving intensity, n (%) |  |
| • Very slight/mild | 12 (5.1) |
| • Mild | 35 (14.8) |
| • Moderate | 98 (41.4) |
| • Severe | 32 (13.5) |
| • Very severe urge | 5 (2.1) |

\*Food noise was defined as unwanted, distracting, and hard-to-stop thoughts about food. Food cravings were defined as a strong urge to eat a specific food or food type that is difficult to resist. Appetite scores range from 0 to 100; higher scores indicate greater hunger, satiety, or fullness, respectively.
N, number of participants with evaluable data; n, number of participants; SD, standard deviation.

### Food cravings prevalence and its intensity

Among participants with evaluable data, the proportion reporting food cravings at least once during a weekly dosing cycle was 52.3% in Week 1 (112/214), 45.5% in Week 2 (95/209), 40.1% in Week 3 (83/207), 41.6% in Week 4 (84/202), 34.9% in Week 5 (65/186), and 31.0% in Week 6 (48/155) (**Figure 3A and Supplementary Figure S2**). Food cravings were reported by 182/237 participants (76.8%) at least once during the 6-week observation period. Across the 6 week observation period, moderate craving was the most frequently reported craving intensity (98/237 [41.4%]), while severe or very severe intensity was reported by 37/237 participants (15.6%). The proportion reporting cravings during each weekly dosing cycle was numerically lower in Week 6 than in Week 1, with a small increase between Weeks 3 and 4; these weekly values are descriptive.

**Figure 3.**
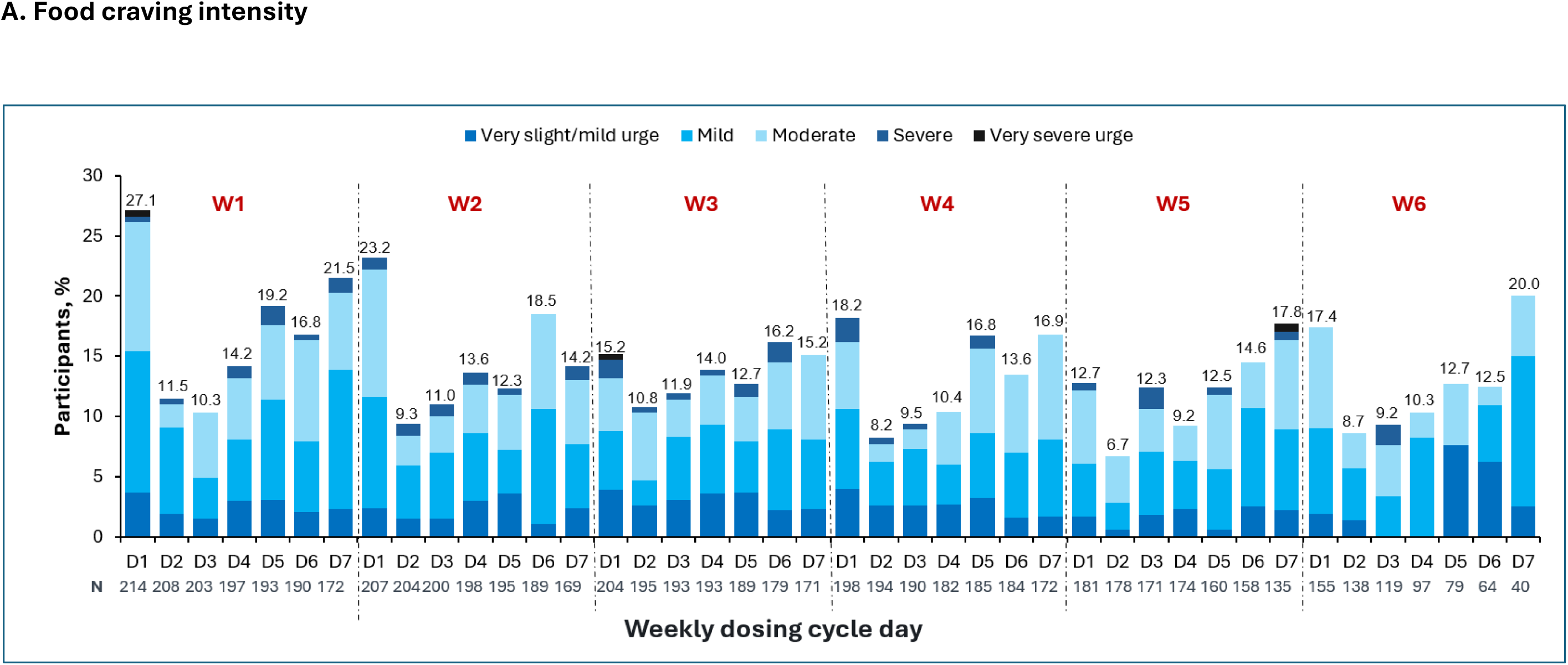
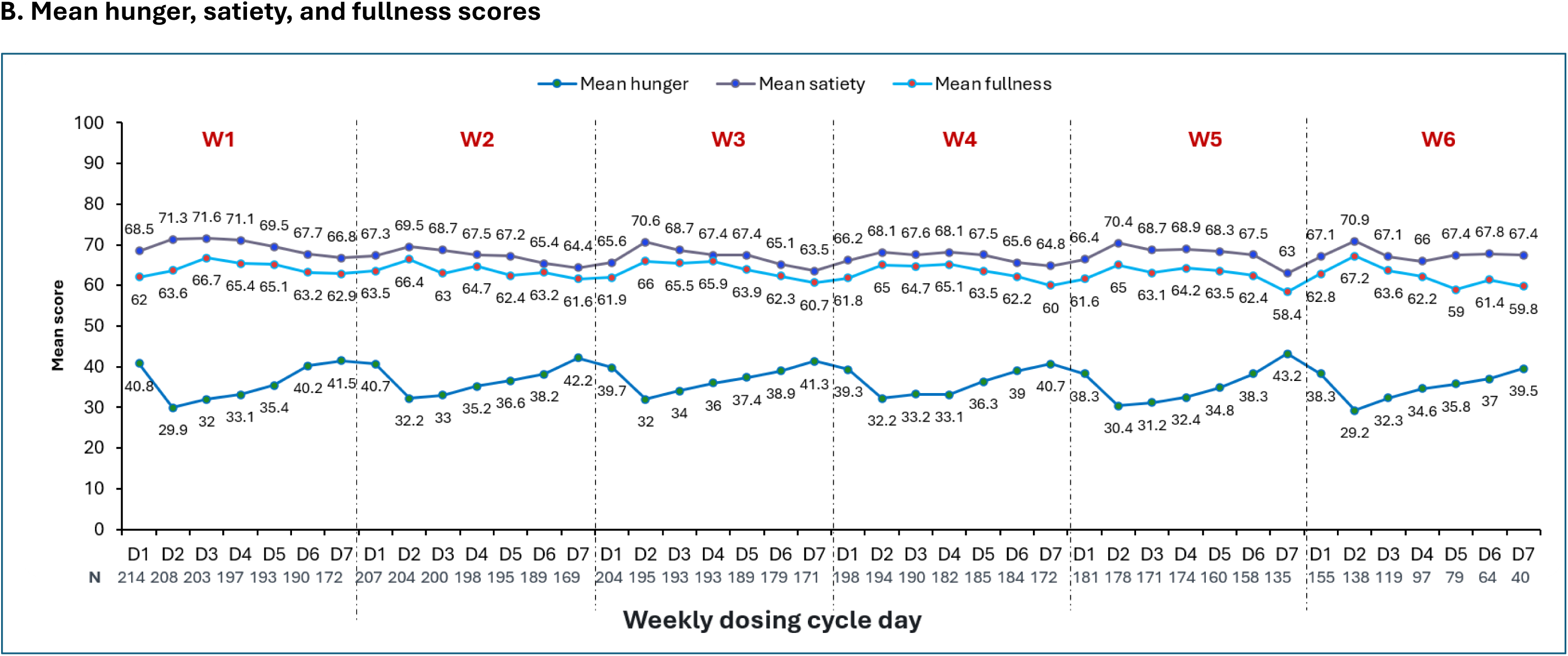
Daily food-related behavior reports by day of the weekly dosing cycle (Primary analysis set) A. Food craving intensity. Stacked bars show the distribution of reported food craving intensity among participants with evaluable craving data for the specified dosing-cycle day. Craving intensity was recorded only when food cravings were reported. N indicates the number of participants with evaluable data for that day. Food cravings were defined for participants as a strong urge to eat a specific food or food type that was difficult to resist. Each dosing cycle began on a participant-reported dose date and included up to the next 7 days or the next reported dose, whichever occurred first. Participants reporting consecutive dosing days were excluded from cycle-aligned analyses. D, day; W, week. Points show mean hunger, satiety, and fullness scores on 0–100 scales among participants with evaluable scores for the specified dosing-cycle day. N indicates the number of participants with evaluable scores for that day. Higher scores indicate greater hunger, satiety, or fullness, respectively. Each dosing cycle began on a participant-reported dose date and included up to the next 7 days or the next reported dose, whichever occurred first. Participants reporting consecutive dosing days were excluded from cycle-aligned analyses. D, day; W, week

### Hunger, satiety, and fullness

Mean hunger scores were lower on Day 2 than on Day 1 in each weekly dosing cycle, then increased gradually through Day 7. Across the six cycles, mean hunger scores (0-100) ranged from 38.3 to 40.8 on Day 1, 29.2 to 32.2 on Day 2, and 39.5 to 43.2 on Day 7.

Mean satiety scores ranged from 65.6 to 68.5, 68.1 to 71.3, and 63.0 to 67.4 on Days 1, 2, and 7, respectively; corresponding fullness-score ranges were 61.6–63.5, 63.6–67.2, and 58.4–62.9 (**Figure 3B and Supplementary Figure S3**). Higher scores indicate greater hunger, satiety, or fullness. Over 6 weeks, median participant-level (n=237) daily scores (Ǫ1–Ǫ3) were 37.0 (29.4–45.8) for hunger, 67.4 (56.5–75.5) for satiety, and 63.1 (52.4–72.5) for fullness.

The patterns observed in Cohorts A and B were generally similar across food noise, food cravings, hunger, satiety, and fullness. Cohort-specific results are presented in **Supplementary Figures S1–S3**.

### Variation in dose interval

Of 242 participants with at least one post-enrollment eDiary assessment, 101 reported at least one interval outside the anticipated 7-day schedule. Among 73 contactable participants, 36 reported erroneous eDiary entries that were corrected post-hoc, and 37 confirmed actual dose-timing variability. Among the 37 participants with confirmed variability, the most common reasons were healthcare professional direction (n=13), travel (n=6), and self-spacing doses because of rapid weight loss (n=2), five provided no reason. Dose trajectories also varied during follow-up: approximately half of Cohort A maintained an unchanged dose, while approximately 20% of Cohort B underwent dose escalation. The reported dates from 28 participants who were not contactable were retained for the purpose of dosing-cycle analyses, but their intervals were not classified as confirmed variability.

## Discussion

In this prospective, direct-to-participant real-world study, daily eDiary assessments described food noise, food cravings, hunger, satiety, and fullness across six consecutive weekly dosing cycles. Reported food noise, cravings, and hunger varied across days within the dosing interval, while satiety and fullness showed less variation. Unlike previous studies that assessed food-related outcomes at scheduled visits, the results from the current study provides a day-by-day description of participants’ reported experiences during ongoing GLP-1–based therapy.

In the present study, the prevalence of daily food noise was generally highest on dosing day, declining thereafter, and returning toward dosing-day levels by Day 7. This prospective assessment complements the findings from the *Impact on Food Noise after Initiating Semaglutide Treatment: Results from a US Survey* (INFORM) study, which retrospectively assessed perceived changes in food noises after semaglutide initiation. ^10^ In INFORM, the median Food Noise Ǫuestionnaire (FNǪ) score (range, 0–20) was 13 before initiation and 6 after initiation. FNǪ captures five different aspects of food noise: time spent thinking about food, persistent food-related thoughts, their negative effects, perceived uncontrollability, and distraction, with each item on a five-point agreement scale. In contrast, the present study used a single daily yes/no item to record whether participants experienced food noise that day to maximize simplicity, given the daily frequency of assessment. Therefore, this item captures its reported presence, but does not assess its frequency, intensity, or specific effects. The findings therefore offer complementary perspectives and should not be directly compared.

The PERCEPTIONS study, a descriptive survey of adults with overweight or obesity who had recently started tirzepatide, was designed to characterize their treatment experiences. Their mean Eating Behavior and Appetite Ǫuestionnaire (EBAǪ) total score was 67.6 on a 0–100 scale (higher scores indicate more favorable appetite and eating behavior. In addition, 64.1% reported often or always feeling full between meals.^11^ Of note, the survey was completed 4–8 weeks after the first dose by 518 participants.

Food noise and food cravings are related but distinct experiences. Food noise refers to unwanted yet difficult-to-stop thoughts about food, whereas food cravings concern a strong urge to eat a particular food or type of food.^12, 13^ In prior treatment studies, food-related outcomes were generally assessed using measures of appetite, cravings, or control of eating rather than a direct assessment of food noise, such as CoEǪ and, Food Craving Ǫuestionnaire-State and Food Craving Inventory. Taken together, the findings from the current study provide context for temporally more granular and holistic food-related experiences during treatment, with a caveat that qualitative characterization of food noise on a daily basis was not intended for feasibility of daily surveys.^12, 14^

Based on visual inspection, temporal patterns in hunger appeared similar to those observed for food noise and food cravings, whereas satiety and fullness exhibited less pronounced. Although the present study was not designed to investigate underlying mechanisms, these observations raise the possibility that GLP-1–based therapies may exert more pronounced or temporally variable effects on appetitive and motivational drives to eat than on meal-related satiation and fullness. Whether these differences reflect distinct neurobehavioral pathways, differential sensitivity to GLP-1 receptor agonism, or varying durations of pharmacologic effects across domains of food-related experience remains unclear. Future mechanistic studies are warranted to investigate these potential explanations.

Dose timing and dose changes are relevant considerations for interpreting day-relative patterns, and thus, our current study assessed two dosing cohorts. The broadly similar food-related patterns observed in Cohorts A and B may partly reflect overlapping dose trajectories during follow-up. Given that 15% of participants reported dosing intervals outside the window and 28 participants have not fully verified administration timing, the Cohort-based analysis should be interpreted cautiously. In clinical settings, the use of GLP-1-based therapies have rapidly evolved, further facilitated by personal preference, tolerability, and more apparent challenges in insurance coverage in the recent few years. Similarly, in a Danish cohort of once-weekly subcutaneous semaglutide users for chronic weight management, only 10% followed the recommended dose increases every 4 weeks, illustrating that real-world dose escalation may differ from trial schedules.^15^ Slower dose escalation may improve tolerability, although its relationship with food-related experiences was not evaluated in the present study.^16, 17^ In this study, reported reasons for actual variation included healthcare professional direction and travel, highlighting the importance of collecting and verifying administration dates in future research.

### Limitations

This study has several limitations. First, although prospective, its nonrandomized, descriptive design does not establish causality, or differences between therapies. Thus, aggregate daily patterns may not reflect consistent participant-level changes. Second, treatment duration was collected categorically, and factors such as dose changes, dietary intake, and daily routines were not assessed or controlled. Third, the inherent limitations of self-reported nature of daily eDiary assessment include declining eDiary completion rate over time and the limited control between eDiary report and drug administration timing should be noted in interpreting results.

### Conclusion and future perspectives

In this prospective real-world study, patient-reported food noise decreased following weekly GLP-1–based therapy administration and increased toward the end of the dosing interval, a temporal pattern consistent with the proposed pharmacokinetic/pharmacodynamic (PK/PD) profiles of these agents. These findings suggest that temporal fluctuations in food-related experiences may have implications for individualized treatment counseling and management, particularly among individuals with obesity in whom intrusive food-related thoughts are a prominent concern. Future prospective studies across a broader range of GLP-1–based therapies, incorporating more granular assessments of food-related experiences, dosing patterns, and agent-specific PK/PD characteristics, are warranted to determine the clinical relevance of the observed patterns, particularly their potential associations with treatment adherence and long-term clinical outcomes.

## Author contributions

Carel W Le Roux, Abd A Tahrani, Kate Orroth, Bethany Kalich, C Michael Gibson, Deborah Horn, Min-Jung Wang, Suna Avcil, Yu Mi Kang, and Zheng-Rong Li contributed to the conception and design of the study. Stephen Fabiano, C Michael Gibson and Yu Mi Kang contributed to patient data collection. All authors, including Chalak Muhammad, Lam-Ǫuynh Nguyen, and Leandro Boer, contributed to the analysis and interpretation of the data. All authors approved the final manuscript and accept responsibility for the integrity of the work.

## Ethical Approval Disclosure

The trial was conducted in accordance with the Good Clinical Practice: Consolidated Guidance (International Council for Harmonization [ICH] E6) and was approved by an independent ethics committee/institutional review board.

## Competing interests’ disclosures

**CLR:** grants from the Irish Research Council, Science Foundation Ireland, Anabio, and the Health Research Board; serving on advisory boards and speakers panels for Novo Nordisk, Herbalife, GI Dynamics, Eli Lilly, Johnson C Johnson, Glia, Irish Life Health, Boehringer Ingelheim, Currax, Zealand Pharma, Keyron, and Rhythm Pharmaceuticals; provides (unpaid) obesity clinical care in the Beyond BMI clinic (Dublin, Ireland), and is a shareholder in the clinic. **DH:** Institutional research funding from Lilly, Kailera, Novo Nordisk, Pfizer, and Weight Watchers. Serving on advisory boards or speaker panels for Amgen, AstraZeneca, Eli Lilly, Kailera, Novo Nordisk, and Zealand. **SF:** employee of Bridge Access Solutions. BK, CM, KO, LB, MW, SA, LN and Z-RL: employees of Amgen Inc. and may own Amgen stock. **CMG:** consultant for Amgen, grants from SCAD Alliance, Johnson C Johnson, Janssen, and CSL Behring; has received personal fees from Bayer, AstraZeneca (during the conduct of the study), Angel Medical Corporation, CSL Behring, Janssen, Johnson C Johnson, the Boston Clinical Research Institute, the Cardiovascular Research Foundation, Gilead Sciences, WebMD, UpToDate in Cardiovascular Medicine, Somahlution, Boston Scientific, the Duke Clinical Research Institute, Medtelligence, Microport, PERT Consortium, Caladrius Biosciences, CeleCor Therapeutics, Eidos Therapeutics, Microdrop, MD Magazine, the Society for Cardiovascular Angiography and Interventions, Revance Therapeutics, Pfizer, Dyad Medical, CytoSorbents, AMAG Pharmaceuticals, Bristol Myers Squibb, Amarin, PhaseBio, Bioclinica, Inari, MedImmune, PLx Pharma, Merck, Paratek, Anthos Therapeutics, PHRI, SmartMedics, EXCITE International ($0 Received), Novo Nordisk, MedTrace, and the Cardiovascular Clinical Science Foundation; has received nonfinancial support from the Baim Institute; and equity from Absolutys, and Dyad Medical. **YMK:** reports consultancy for Baim Institute for Clinical Research and Bridge Access Solutions; advisory board role for Novo Nordisk Inc.

## Data sharing

Ǫualified medical and scientific researchers may submit requests for de-identified individual participant data, study protocols, statistical analysis plans, and other study documents from completed Amgen clinical trials and observational studies. Requests are subject to Amgen’s data-sharing process and applicable legal and ethical requirements. Details are available at [Amgen Data Sharing Request](https://www.amgen.com/science/clinical-trials/clinical-data-transparency-practices/%20clinical-trial-data-sharing-request).

## Funding

This study was funded by Amgen Inc.

## Data Availability

Qualified medical and scientific researchers may submit requests for de-identified individual participant data, study protocols, statistical analysis plans, and other study documents from completed Amgen clinical trials and observational studies. Requests are subject to Amgen data-sharing process and applicable legal and ethical requirements

https://www.amgen.com/science/clinical-trials/clinical-data-transparency-practices/clinical-trial-data-sharing-request

## Acknowledgments

The authors thank the study participants for taking part in this study. Medical writing support was provided by Aditya Pramod, Amgen India, in accordance with Good Publication Practice (GPP 2022) guidelines.

## Notes

### Author Declarations

The trial was conducted in accordance with the Good Clinical Practice: Consolidated Guidance (International Council for Harmonization [ICH] E6) and was approved by the Advarra Institutional Review Board (Advarra IRB), Columbia, Maryland, USA.

