## Supplemental Table S1 for "Patient-Reported Food Noise, Cravings and Appetite in Adults with Obesity Treated with Weekly Injectable GLP-1 Based Therapies: A Prospective Real-World Study"

**Supplementary Table S1. Participant eligibility criteria**

| **Category** | **Criterion** |
| --- | --- |
| **Inclusion** | Adult aged ≥18 years at screening |
|  | Self-reported initial BMI ≥30 kg/m² before starting GLP-1-based therapy. BMI at study enrollment was not required to be ≥30 kg/m². |
|  | Planned to continue GLP-1-based therapy for the 6-week study period |
|  | Willing to adhere to GLP-1-based therapy during the study |
|  | Willing to provide feedback through daily electronic diaries or surveys |
|  | Met the treatment-status criteria for Cohort A or Cohort B in Supplementary Table S1 |
| **Exclusion** | Use of compounded GLP-1-based treatment |
|  | Use of a non-FDA-approved GLP-1-based treatment dose, such as a microdose |
|  | Use of other obesity or weight-management medications, over-the-counter products, or supplements within 3 months before enrollment, or plans to use them during the study |
|  | Type 1 or type 2 diabetes |
|  | Pregnancy or breastfeeding |
|  | A relevant or serious chronic medical condition, including visual problems or severe mental illness that could prevent participation in study activities |
|  | Bariatric surgery within the previous year or plans to undergo bariatric surgery within the next 3 months |

Eligibility criteria were assessed using participant-reported information; reported information was verified against available electronic medical record data through Health Information Exchange linkage, as applicable. The BMI criterion refers to self-reported BMI before starting GLP-1-based therapy; BMI at study enrollment was not required to be ≥30 kg/m².

BMI, body mass index; EMR, electronic medical record; FDA, US Food and Drug Administration; GLP-1, glucagon-like peptide-1; HIE, Health Information Exchange.
