## Supplemental Table S2 for "Patient-Reported Food Noise, Cravings and Appetite in Adults with Obesity Treated with Weekly Injectable GLP-1 Based Therapies: A Prospective Real-World Study"

**Supplementary Table S2. Summary of cognitive-debriefing findings**

| **Aspect reviewed** | **Summary of feedback** |
| --- | --- |
| **Instructions and navigation** | The feedback log raised the need to clarify how to proceed after a “No” response and how to complete the rating items. It also noted that spacing and presentation could be improved. |
| **Relevance and comprehensiveness** | The log noted potential conceptual overlap between satiety and fullness items and uncertainty about whether their wording and anchors distinguish the concepts. |
| **Response options and scale usability** | The log questioned whether “Not at all” was appropriate for follow-up items shown after a “Yes” response. It also raised whether numeric labels or other descriptors would help with rating scales. |
| **Suggested wording changes not supported** | Suggestions were made to remove wording, considered redundant. |

Cognitive debriefing was a qualitative assessment of the participant-facing materials and eDiary items; it did not constitute psychometric validation of the ePRO.

eDiary, electronic diary.
