## Supplemental Table S3 for "Patient-Reported Food Noise, Cravings and Appetite in Adults with Obesity Treated with Weekly Injectable GLP-1 Based Therapies: A Prospective Real-World Study"

**Supplementary Table S3. Food-related items and response options from the daily eDiary**

| **Outcome** | **eDiary item** | **Response options or scale** |
| --- | --- | --- |
| **Food noise presence** | “In the last 24 hours, did you have food noise? Food noise is unwanted, distracting, and hard-to-stop thoughts about food.” | Yes / No |
| **Negative impact of food-related experiences** | “In the last 24 hours, my thoughts about food have had negative effects on me and/or my life.” | Yes / No |
| **Food cravings** | “In the last 24 hours, did you have food cravings? Food cravings are a strong urge to eat a specific food or food type that is difficult to resist.” | Yes / No |
| **Craving intensity** | “How strong were your food cravings in the last 24 hours?” | Very slight/mild urge; mild; moderate; severe; very severe urge |
| **Hunger** | “How hungry did you feel in the last 24 hours? Hunger means the physical feeling your body needs food, such as a growling stomach or feeling like you need to eat.” | 0 (“I am not hungry at all”) to 100 (“I have never been hungrier”) |
| **Satiety** | “How satisfied did you feel in the last 24 hours? Satiety is feeling content or pleased after eating and whether your meals felt satisfying and met your needs.” | 0 (“I am not satisfied at all”) to 100 (“I have never been more satisfied”) |
| **Fullness** | “How full did you feel in the last 24 hours? Fullness means the physical feeling of your stomach being filled with food, such as feeling stuffed or stretched after eating.” | 0 (“I am not full at all”) to 100 (“I have never been fuller”) |

All items refer to the preceding 24 hours. The craving-intensity item was completed only when food cravings were reported.

eDiary, electronic diary.
