## Supplemental Table S4 for "Patient-Reported Food Noise, Cravings and Appetite in Adults with Obesity Treated with Weekly Injectable GLP-1 Based Therapies: A Prospective Real-World Study"

**Supplementary Table S4. Negative impact of food-related thoughts by cohort**

| **Analysis interval (Weekly dosing cycle)** | **Cohort A, n/N (%)** | **Cohort B, n/N (%)** | **Total, n/N (%)** |
| --- | --- | --- | --- |
| **Week 1** | 18/109 (16.5) | 15/105 (14.3) | 33/214 (15.4) |
| **Week 2** | 12/106 (11.3) | 15/103 (14.6) | 27/209 (12.9) |
| **Week 3** | 14/105 (13.3) | 10/102 (9.8) | 24/207 (11.6) |
| **Week 4** | 11/103 (10.7) | 13/99 (13.1) | 24/202 (11.9) |
| **Week 5** | 12/93 (12.9) | 12/93 (12.9) | 24/186 (12.9) |
| **Week 6** | 5/79 (6.3) | 8/76 (10.5) | 13/155 (8.4) |
| **Overall 6-week period** | 41/114 (36.0) | 44/123 (35.8) | 85/237 (35.9) |

Data are shown as n/N (%), where *n* is the number of participants reporting a negative impact of food-related thoughts and *N* is the number with evaluable data for the specified interval. A negative impact was defined as a “Yes” response to the daily eDiary item: “In the last 24 hours, my thoughts about food have had negative effects on me and/or my life.” Participants reporting consecutive dosing were excluded from weekly dosing-cycle summaries and included in the overall 6-week summary. Cohort A comprised participants who reported starting GLP-1 therapy 1 to <12 weeks before enrollment; Cohort B comprised participants who reported a stable dose for at least 12 weeks before enrollment. GLP-1, glucagon-like peptide-1; eDiary, electronic diary.
