## Supplemental Figure S1 for "Patient-Reported Food Noise, Cravings and Appetite in Adults with Obesity Treated with Weekly Injectable GLP-1 Based Therapies: A Prospective Real-World Study"

**Supplementary Figure S1. Daily food noise reports by day of the weekly dosing cycle and cohort**

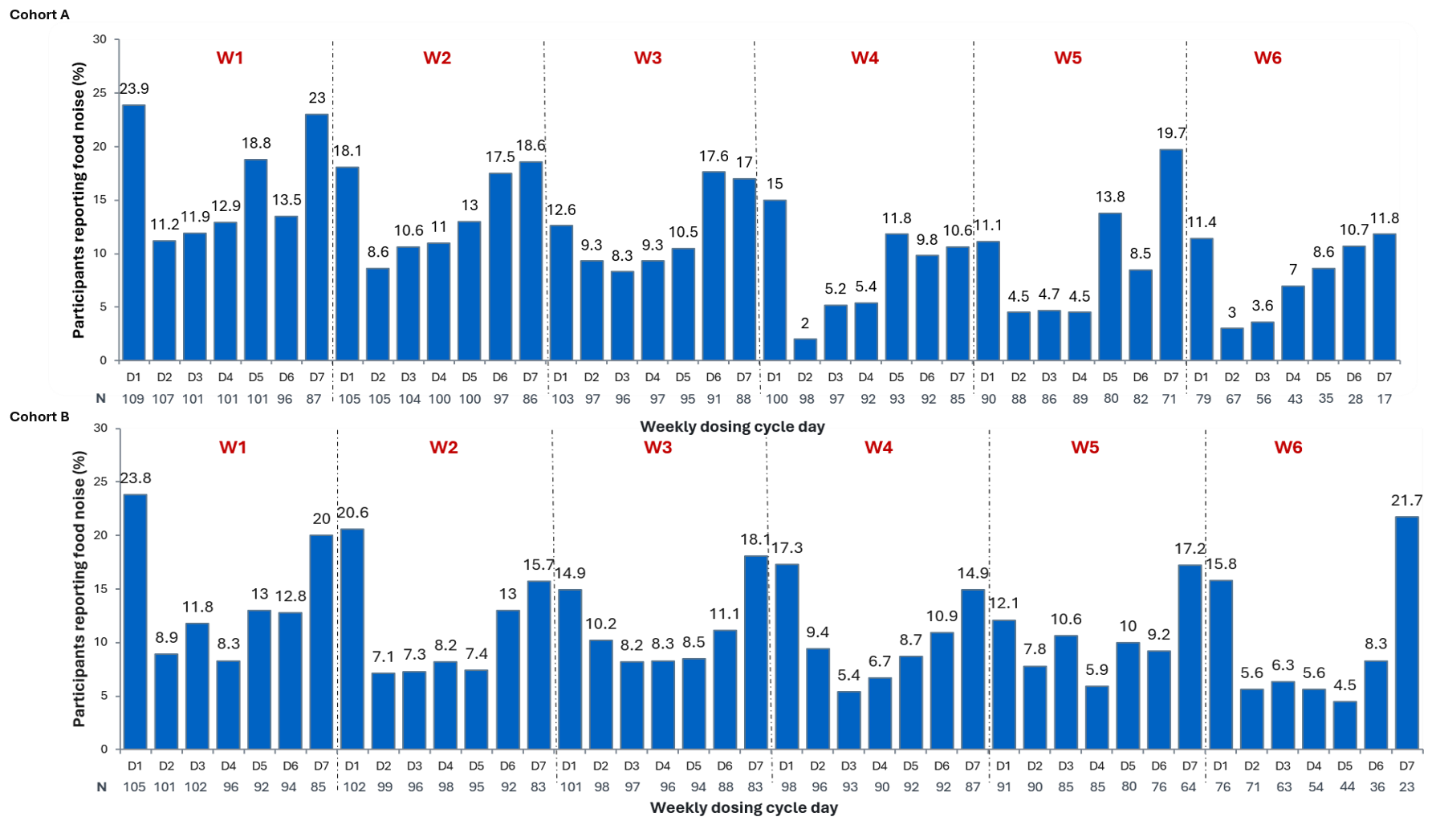

Values are percentages of participants with evaluable data for the specified outcome and dosing-cycle day. N indicates the number of participants with evaluable data for that day. Participants reporting consecutive dosing days were excluded from cycle-aligned analyses. Cohort A included participants who had received therapy for 1 to <12 weeks before enrollment and were not yet on a stable dose. Cohort B included participants who had received a stable dose for ≥12 weeks before enrollment, with no planned change in dose, dosing frequency, or product during the study. D, day; W, week
