## Supplemental Figure S3 for "Patient-Reported Food Noise, Cravings and Appetite in Adults with Obesity Treated with Weekly Injectable GLP-1 Based Therapies: A Prospective Real-World Study"

**Supplementary Figure S3: Mean hunger, satiety, and fullness scores by day of the weekly dosing cycle and cohort**

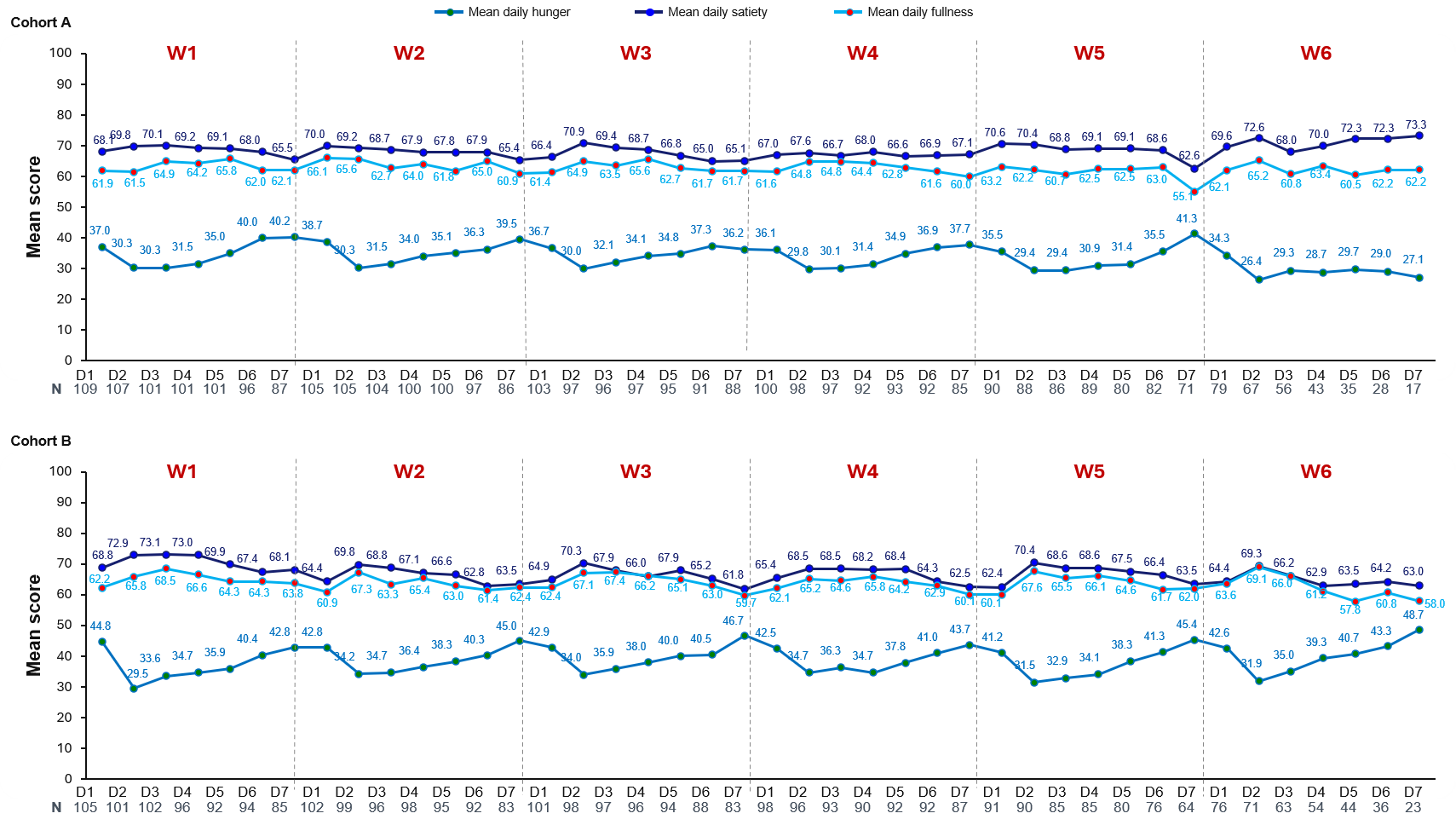

Points show mean hunger, satiety, and fullness scores on 0–100 scales among participants with evaluable scores for the specified dosing-cycle day. N indicates the number of participants with evaluable scores for that day. Higher scores indicate greater hunger, satiety, or fullness, respectively. Participants reporting consecutive dosing days were excluded from cycle-aligned analyses. Cohort A included participants who had received therapy for 1 to <12 weeks before enrollment and were not yet on a stable dose. Cohort B included participants who had received a stable dose for ≥12 weeks before enrollment, with no planned change in dose, dosing frequency, or product during the study. D, day; W, week
